# Were Bangladesh’s Growth Monitoring and Promotion (GMP) programmes resilient during a health system crisis? A qualitative study

**DOI:** 10.64898/2026.01.22.26344566

**Authors:** Muttaquina Hossain, Md. Fakhar Uddin, Tiia Haapaniemi, Tahmeed Ahmed, Ulla Ashorn, Per Ashorn

## Abstract

**Introduction:** In Bangladesh, growth monitoring and promotion (GMP) is delivered through both facility-based (FGMP) and non-government organisation (NGO)-supported home-plus-facility approaches (HGMP+FGMP). The COVID-19 pandemic disrupted health services, but little is known about the resilience of GMP delivery in rural community clinics (CCs) during this health system crisis. This study examined facilitators and barriers to GMP utilisation and implementation before and during COVID-19.

**Methods:** We conducted a longitudinal qualitative study in six rural sub-districts of Mymensingh at two timepoints before and after the lockdown of the COVID-19 crisis. We interviewed 84 participants, including government officials, CC providers, NGO managers, and caregivers of children under two, and indirectly observed GMP service provision at 23 CCs before lockdown. Data were thematically analyzed using the WHO’s health systems building blocks framework to examine resiliency during a health system crisis.

**Results:** Caregivers’ awareness and engagement in GMP were low across both programmes and timepoints. Most CC providers and caregivers equated GMP with weight measurement for sick children alone. Length/height measurement and counselling were rarely provided. HGMP+FGMP improved access and uptake before the pandemic through home visits and immunization programme integration. Common barriers included workforce shortages, high workloads, weak government–NGO coordination, limited logistics and poor infrastructure, and inadequate supervision. The COVID-19 pandemic exacerbated these challenges, leading to partial suspension of FGMP and complete suspension of home-based services. Staff redeployment, supply chain disruptions, and caregiver disengagement were reported during the pandemic. Both programmes lacked health system–level guidance for crisis response.

**Conclusions:** GMP programmes in rural Bangladesh exhibited limited resilience during a health system shock. Integrated HGMP+FGMP models can enhance access and engagement, but they need sustained institutional support, workforce capacity, caregiver communication, and resiliency and preparedness policies to maintain programme continuity during crises.Strengthening these areas can safeguard essential child nutrition services in future shocks.

**Key messages:** *What is already known:*

- Like in other settings globally, health services in Bangladesh face persistent implementation challenges and are disrupted during health system shocks, such as the COVID-19 pandemic.

*What this study adds:*

- Growth Monitoring and Promotion (GMP) programmes in Bangladesh lacked resilience, with pre-existing health system weaknesses limiting their ability to adapt and maintain service delivery during a crisis.

*How this study might affect research, practice or policy:*

- Strengthening GMP programmes requires integrating emergency preparedness, improving caregiver communication, building provider capacity, and ensuring sustainable resources and system-level support.
- Embedding resilience principles into GMP programme design, including community outreach, supervision, workforce support, and contingency planning, can help safeguard essential child nutrition services during future health system crises.

## Introduction

Growth Monitoring and Promotion (GMP) is a key component of child health programmes in low- and middle-income countries (LMICs), aimed at detecting growth faltering among children under five and providing timely counselling and referrals to prevent or manage illness^1^ ^2^. In Bangladesh, GMP is primarily delivered through government facility-based platforms, including community clinics (CCs)^3^, but have low coverage and utilisation^4^. To improve access and utilization, a non-governmental organization (NGO) piloted a home-based GMP (HGMP) initiative in selected rural sub-districts, delivered by trained frontline workers at households during 2018-2020. While HGMP achieved higher uptake than facility-based GMP (FGMP) before the pandemic, in a 2020 evaluation, overall utilisation remained low (40%), and both models fell short of improving child growth and feeding practices^5^ ^6^.

Common barriers to GMP delivery in LMICs include inadequate training and supervision, limited logistics and equipment, low caregiver awareness, poor data recording, and weak integration of counselling ^7–11^. Facilitators such as structured capacity-building, supportive supervision, integration of nutrition education with growth assessment, integration with immunization programme, home-based delivery, and digital growth tracking have shown promise in some LMICs^6^ ^12^ ^13^, but are often inconsistently applied. The COVID-19 pandemic created a major global health system shock, disrupting essential preventive child health services and risked reversing gains in malnutrition reduction. Studies from LMICs have documented the widespread global impact of COVID-19 on maternal and child health services^14^ ^15^. In Bangladesh, routine GMP services at community clinics were halted, and NGO-led HGMP activities were suspended and not resumed. However, there is limited evidence on how GMP delivery mechanisms in Bangladesh were affected, particularly from the perspectives of caregivers, frontline providers, and policymakers. To our knowledge, no studies have systematically examined how GMP programmes in Bangladesh adapted or failed to adapt during a major health system shock, nor analysed resilience from a multi-stakeholder, health systems perspective. This study used in-depth, qualitative methods to examine whether and how Bangladesh’s GMP programme demonstrated resilience during a health system crisis, analyzing system-level facilitators and barriers before and after the COVID-19 lockdown. Specifically, it explored institutional and programmatic challenges, stakeholder perceptions, and caregiver experiences. This study sought to determine whether Bangladesh’s GMP programmes were resilient to the COVID-19 health system shock, and to identify the health system factors that facilitated or hindered service continuity and adaptation. The findings aim to provide actionable insights to strengthen GMP delivery mechanisms and safeguard essential child health services amid crises in Bangladesh, and draw lessons for other settings.

## Methods

### Study Design, setting and timeline

We conducted this qualitative study as a part of a quasi-experimental evaluation of two GMP delivery models between August 2019 and December 2020^5^. It took place in six rural sub-districts of Mymensingh division, a region 120 km north of Bangladesh’s capital, Dhaka, with a population of 6 million approx. During the study period, two GMP models were implemented: (1) government-led, facility-based GMP (FGMP) and (2) NGO-led, home-based GMP (HGMP), which was delivered alongside FGMP (HGMP+FGMP). Each model was allocated to three sub-districts. The parent evaluation assessed the effectiveness of government-led, facility-based GMP (FGMP) versus NGO-led, home-based GMP delivered alongside FGMP (HGMP+FGMP) on child growth, feeding practices, and service utilization^4^ ^5^. Our in-depth qualitative study complemented this by exploring stakeholder perspectives, caregiver experiences, and health system-level factors influencing the continuity, adaptation, and resilience of GMP services during the COVID-19 pandemic.

During the study period, the Government of Bangladesh (GoB) imposed a nationwide lockdown (March –May, 2020) to curb COVID-19 infections, which disrupted routine health and nutrition services. We applied a longitudinal qualitative design, conducted interviews at baseline (pre-lockdown, July–December 2019) and endline (post-lockdown, October–December 2020). This design enabled an assessment of health system resilience by examining changes in the continuity, disruption, and adaptation of GMP services before and during a major health system shock. We could not collect data during the lockdown period due to movement restrictions and ethical considerations for participants’ and researchers’ safety.

### Components of the GMP programme

In Bangladesh, the GoB’s FGMP programme targets quarterly growth monitoring for all children under five years of age in all primary healthcare facilities, including CCs. Each CC delivered FGMP services through Community Health Care Providers (CHCPs), Health Assistants (HAs), and Family Welfare Assistants (FWAs). The CC providers were responsible for measuring child weight, length/height, and mid-upper arm circumference (MUAC); plotting measurements on GMP cards; providing counselling based on growth status; and referring children identified as undernourished^3^. CHCPs received a five-day basic nutrition training that included eight hours on GMP procedures, though most received this training 2-5 years prior to the study with no refresher training. The government supplied equipment and materials for FGMP delivery. However, FGMP lacked a community-based child tracking or unique identification system to support longitudinal follow-up.

In selected sub-districts, an NGO delivered home-based GMP (HGMP) services to complement FGMP. The NGO conducted household visits, coordinated GMP activities with CCs and EPI clinics, supplied GMP-specific behaviour change communication materials, and established a community-based child identification and tracking system. Monthly coordination meetings were held between NGO staff and CC providers. These additional components were designed to enhance service reach, continuity, and follow-up, features relevant to health system resilience.

### Study Participants

We conducted 84 interviews across baseline (n=48) and endline (n=36). Participants included government stakeholders (e.g., national nutrition programme directors and managers), community clinic workers, NGO stakeholders and field workers, and mothers/caregivers of children under two years of age (Appendix). We applied purposive and snowball sampling to select participants with diverse roles in GMP implementation and utilization. We recruited caregivers from the parent quasi-experimental study sampling frame^5^ to capture variation in programme exposure, geographic location, and experiences of GMP service availability before and during COVID-19 lockdown.

### Data Collection

Our data collection team consisted of one moderator (male) and two interviewers (1 male and 1 female) fluent in Bangla and trained in qualitative methods. We used semi-structured guides, validated through translation and back-translation, that explored GMP availability, service continuity, access barriers, facilitators, and adaptations before and during COVID-19.

Interviews lasted 30–60 minutes. We conducted interviews at a location convenient for the participants, ensuring privacy. We revisited participants when we needed clarification or additional information with prior consent. The data collectors continued interviews until they reached redundancy in information and thematic data saturation. The reduced sample at endline (36 vs. 48 interviews) reflected that many themes were reinforced rather than new. COVID-19 limited access to some participants, particularly NGO programme managers and field workers engaged in emergency response activities, or were discontinued from the HGMP programme. We assessed saturation through team consensus, confirming that no new codes or themes appeared in the final three interviews within each participant category.

### Conceptual framework

To examine how Bangladesh’s GMP programmes responded to the COVID-19 health system shock, we applied a health systems resilience conceptual framework. Health system resilience to shock is defined as the capacity of a system to absorb, adapt, and transform in response to shocks while maintaining core functions^16^ ^17^. We drew on Thomas et al.,^18^ which frames resilience as the ability to prepare for, manage (absorb, adapt, transform), and learn from shocks. We focused on the *shock impact and management stage*, capturing service disruption, adaptation, and resource reorganisation during crises. To operationalise the examination of health system resilience, we aligned the framework with the WHO Health Systems Building Blocks^19^, examining GMP service delivery, workforce, equipment and infrastructure, health information systems, leadership and governance, and financing (Table 1). This framework guided data collection, coding, and interpretation of system-level facilitators and barriers affecting GMP resilience before and during COVID-19.

**Table 1.** GMP service resilience defined as per WHO building blocks.

| WHO building blocks | Definition of Resilience variables examined in the study |
| --- | --- |
| GMP service delivery | Continuity of GMP services, service suspension, adaptation of delivery platforms (home visits, EPI integration, CC courtyard), caregiver access during COVID-19 |
| Health workforce | CC and NGO GMP service provider availability, redeployment during COVID-19, workload pressures affecting GMP continuity |
| Equipment & infrastructure | Availability and functionality of anthropometric equipment required to continue GMP during disruption, CC infrastructure and space for GMP service continuation |
| Health information | Continuity of child tracking in community, data use for follow-up, |

| **WHO building blocks** | **Definition of Resilience variables examined in the study** |
| --- | --- |
| systems | supervision and feedback during the shock |
| Leadership & governance | Presence or absence of operational GMP guidance, coordination mechanisms, and crisis-response decision-making, emergency/shock resilience policy and guidelines for GMP programme |
| Financing | Availability or diversion of resources affecting GMP continuity during COVID-19 |

CC: Community clinic; COVID-19: coronavirus disease 2019; EPI: Expanded Programme on Immunization; GMP: growth monitoring and promotion; NGO: non-governmental organization.

### Data Analysis

With consent, the data collectors audio-recorded all interviews, transcribed them verbatim in Bangla, translated them into English, and repeatedly reviewed the transcripts to ensure accuracy and familiarisation. Two researchers (MH and FU) trained in qualitative methodology conducted a thematic analysis using qualitative content analysis^20^. We applied a combined deductive and inductive coding approach. We developed an initial codebook deductively from the study objectives and the WHO Health Systems Building Blocks framework^19^. We then expanded it inductively based on participants’ narratives, including caregiving practices, household dynamics, and responses to COVID-19 related service disruptions.

We coded transcripts line by line and supported the analysis with analytic memos that documented emerging interpretations and decisions. We organised coding using structured analytic worksheets. We created separate worksheets for each respondent group (caregivers, community clinic healthcare providers, and NGO managers) and for each time point (before and after the COVID-19 lockdown). Within each worksheet, we organised data by thematic domains and subthemes relevant to service continuity, access, quality, and adaptation during the pandemic. We visually distinguished inductive and deductive codes to facilitate comparison across groups and timelines and to identify patterns related to health system resilience.

We independently coded 20% of transcripts to check intercoder reliability (85%) and resolved discrepancies by consensus to ensure rigor. We resolved coding discrepancies through discussion and consensus, then applied the revised codebook to all transcripts. We summarised coded data into analytical matrices organised by WHO health system building block, respondent type, and time period. We used constant comparison techniques to examine similarities and differences across programme models and over baseline and endline. We triangulated findings across interviews, facility observations, and programme documents to strengthen credibility and trustworthiness. We enhanced transferability by providing detailed descriptions of the study context and participant characteristics^21^ ^22^. We achieved confirmability through reflexive discussions between analysts and by maintaining an audit trail of analytic decisions. We present direct quotations from anonymised participants in the results to illustrate key themes and to retain participants’ voices, while situating findings within the health system framework. We organised the final analytical framework around the six WHO Health Systems Building Blocks to interpret system-level facilitators and barriers to GMP service delivery and resilience during the COVID-19 health system shock. We treated caregiver perspectives as cross-cutting insights that illustrate how health system factors shaped service access, continuity, and utilisation across programme models and time points. We followed the COREQ (Consolidated Criteria for Reporting Qualitative Research) checklist to ensure transparency and methodological rigour^23^ (Appendix).

### Ethics

The Institutional Review Board of icddr,b approved the study (Protocol ID: PR-17123, version 1.02, 8 March 2018). We obtained written informed consent from all participants, after clearly explaining the study’s aims, procedures, risks, and benefits. We removed personal identifiers from transcripts and restricted access to sensitive data to principal investigators. The parent evaluation was registered prospectively at ClinicalTrials.gov in January 2019 (ID: NCT03824756).

## Results

### Participants, data sources, and service context

We conducted 84 in-depth interviews with 77 unique participants across baseline (n=48) and endline (n=36), including government stakeholders (n=8), CC healthcare providers (n=31), NGO programme managers and field workers (n=10), and caregivers (n=35) from both HGMP+FGMP and FGMP-only areas (appendix). We observed GMP activities at 23 CCs and 5 EPI clinics at baseline. Endline data collection focused on experiences during and after the COVID-19 lockdown from all respondents without CC observations. Results are organised according to resilience-relevant domains derived from the WHO Health System Building Blocks framework, focusing on service continuity, workforce availability, infrastructure and supplies, health information systems, and leadership and financing across baseline and post-COVID-19 endline periods.

The primary caregivers were mothers of under-2 children in both areas. They were typically housewives, aged between 24-26 years with 7-9 years of education. Fathers were mostly service holders or day labourers. The monthly household incomes ranged from 9,571 to 13,000 BDT (78 to 106 US dollars), with slightly higher income at FGMP+HGMP areas at both time points. The family sizes were comparable in both areas at both time points. CC providers were aged between 32-34 years, had 15-17 years of education, and 8-9 years of work experience. CHCPs gender distribution varied by site: HGMP+FGMP areas had more balanced male–female representation, whereas FGMP-only sites were predominantly female (supplementary table 1-2).

### Demand-side factors affecting service continuity

In HGMP+FGMP areas, caregivers received growth monitoring of their children through NGO home visits, EPI clinics, or CCs at baseline. Five of 11 caregivers (45%) reported receiving GMP cards and having their child’s weight measured, and four (36%) could interpret a growth chart (Table 2). None of the caregivers reported having ever received GMP-specific counselling, but they reported having received general counselling on child feeding. At endline, after the lockdown and following cessation of NGO activities, none of the caregivers reported receiving GMP cards, home visits, or weight or length measurement. Awareness of where to obtain GMP services declined from 45% at baseline to 0% at endline. Caregivers reported uncertainty about alternative points of care:

**Table 2.**
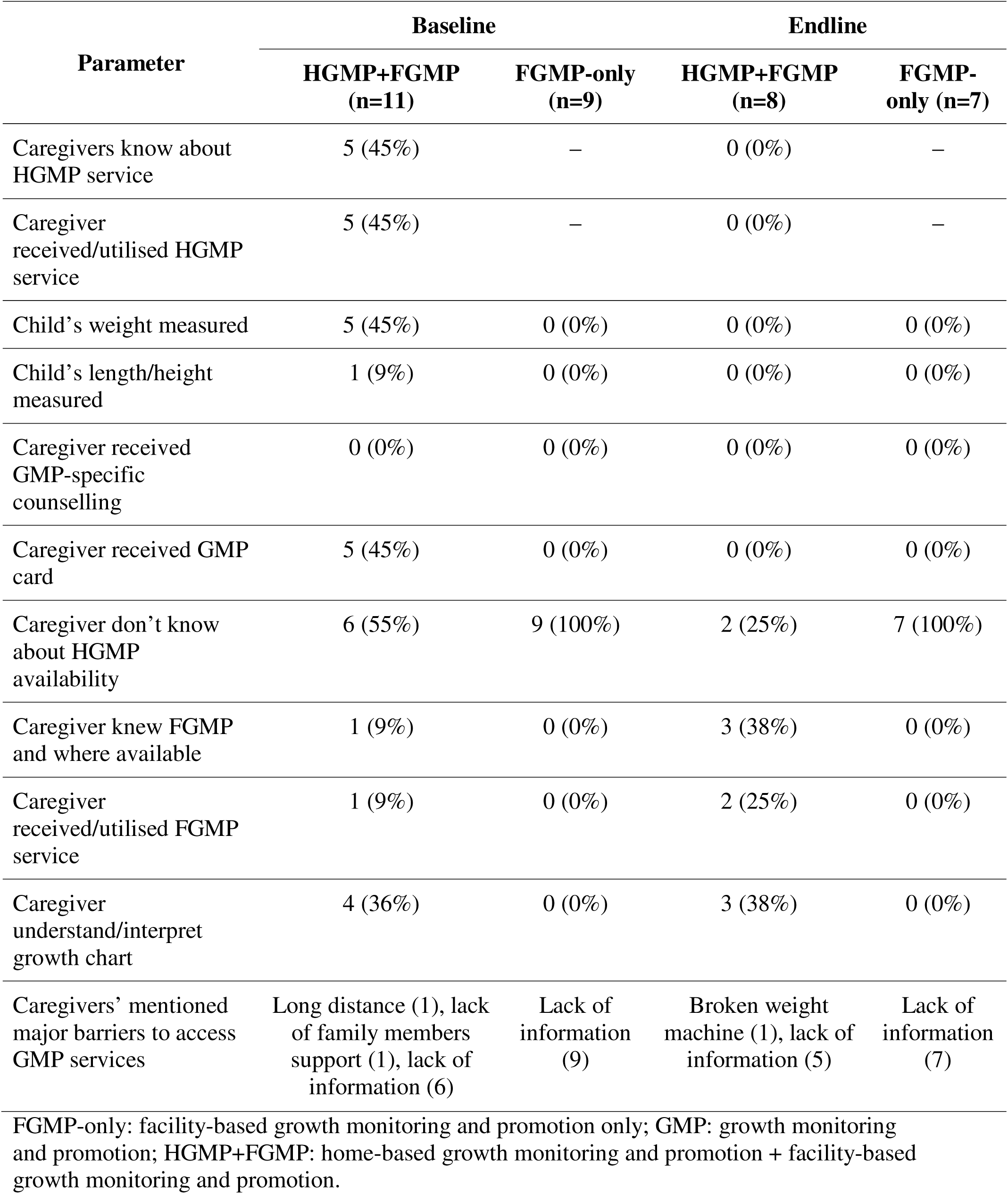
Caregivers’ knowledge and utilization of GMP services.

“*None of us knew where to take the child after the NGO programme stopped*.” (Caregiver, HGMP+FGMP, endline).

In FGMP-only areas, no caregivers reported receiving weight or length measurement, GMP cards, or GMP-specific counselling at either time point. At both baseline and endline, all caregivers (100%) reported not knowing about GMP service availability at CCs (Table 2):

“*No one ever told me about this (GMP) service. I thought the clinic (CC) was only for medicine.”* (Caregiver, FGMP, endline).

### GMP programme resilience during health system shock

Before the COVID-19 lockdown, GMP in HGMP+FGMP areas was delivered through multiple channels, including home visits, EPI sessions, and CCs (Table 3). All eight NGO field workers reported that home visits stopped completely from March 2020 and had not resumed by December 2020. An NGO field worker stated that:

**Table 3.**
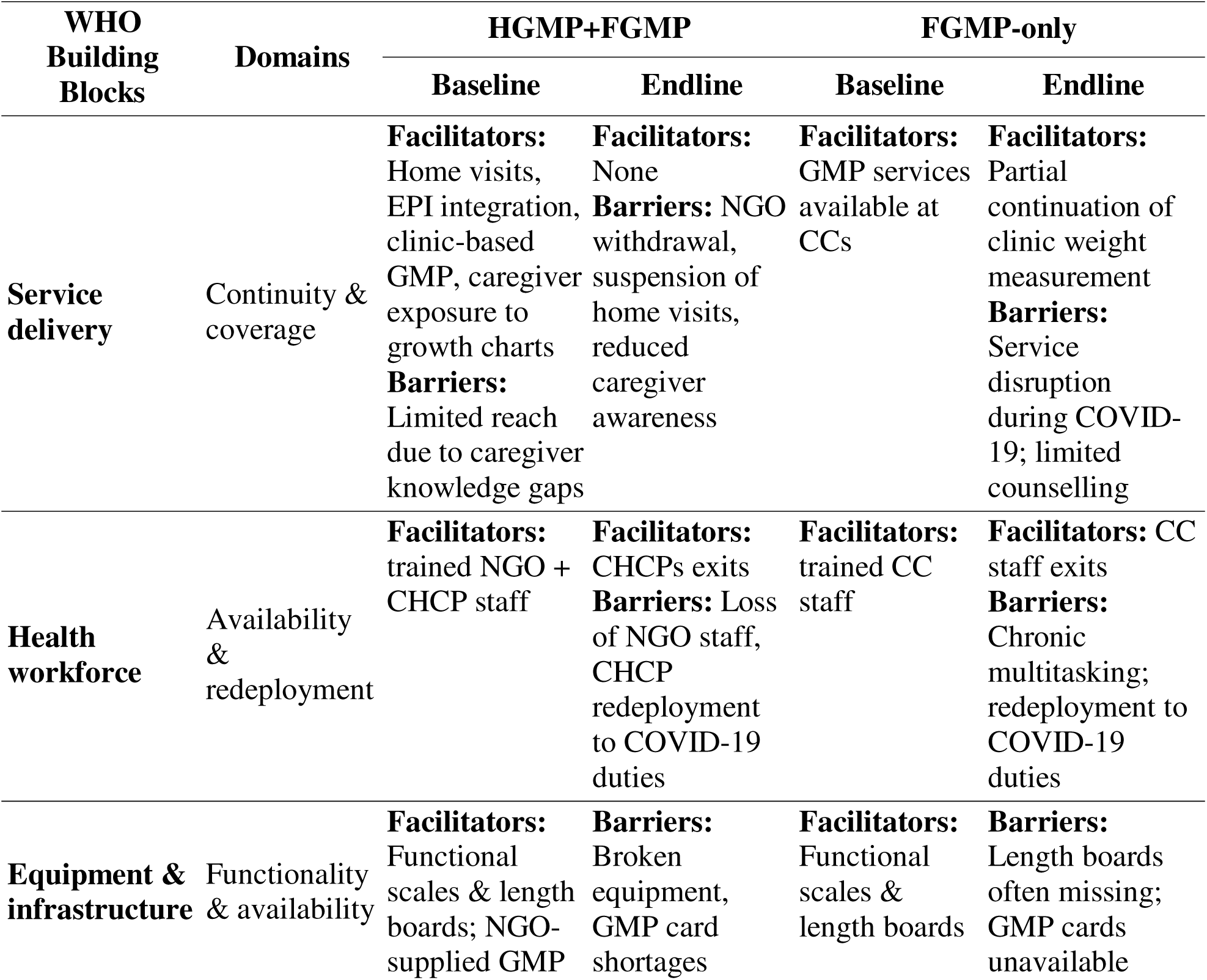
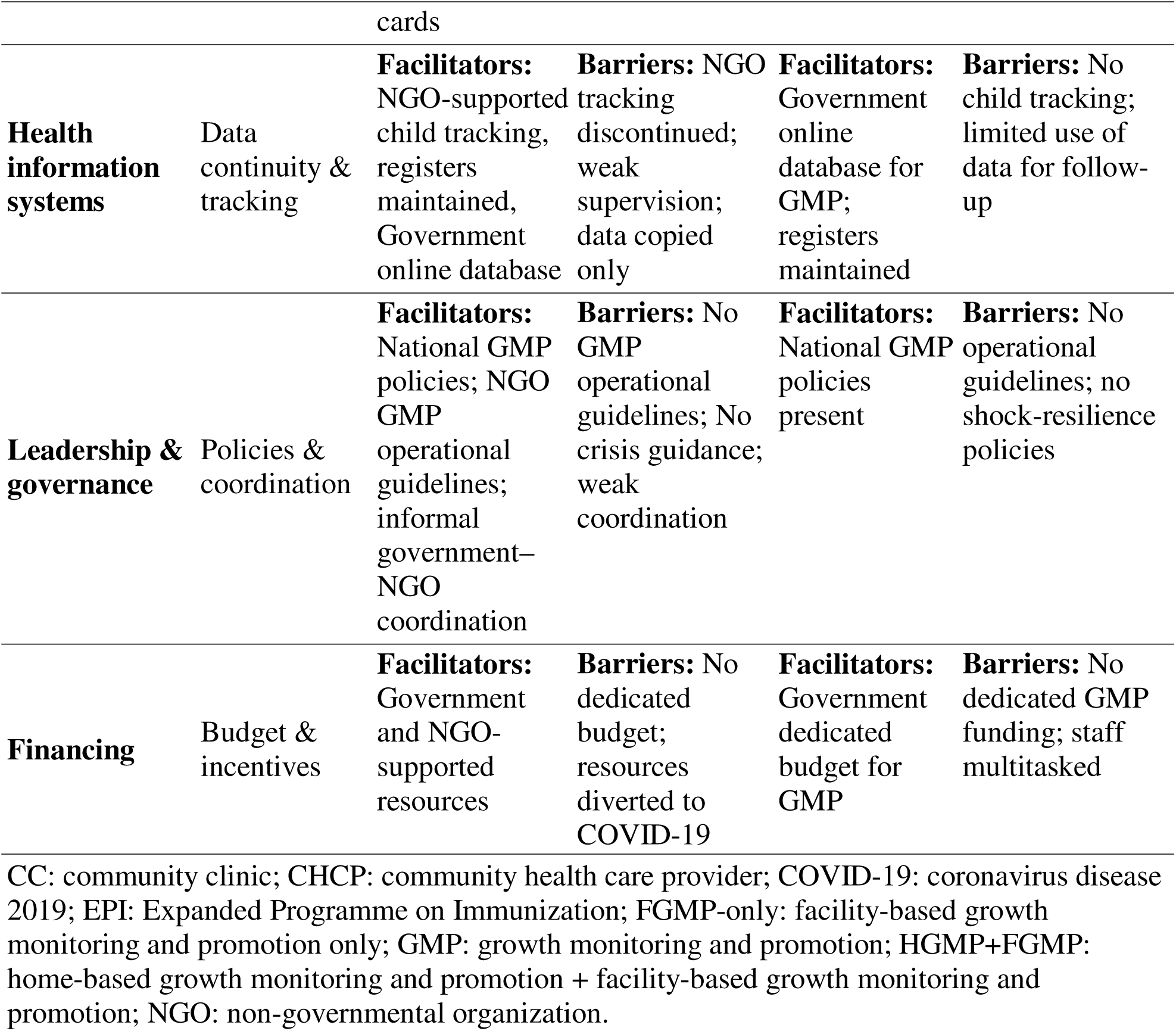
Health system–level barriers and facilitators affecting GMP programmes resilience before and during COVID-19.

“*We visited households regularly before lockdown; after lockdown, we only focused on COVID work*.” (NGO Field Worker, HGMP+FGMP, endline).

In both areas, CC providers reported that routine growth monitoring was suspended during lockdown, with services limited mainly to sick-child care. Five of seven CHCPs reported that no routine GMP was conducted during this period. By endline, caregivers in both areas reported that most children had not received recent weight or length measurement and were unsure whether GMP services were available. In FGMP-only areas, GMP delivery was limited to clinic-based weight measurement before COVID-19 and remained clinic-dependent during lockdown, with partial disruption reported by CC providers (Table 3).

### Health workforce availability and redeployment

At baseline, CHCPs in both models mentioned conducting routine GMP activities alongside other responsibilities at CCs. In HGMP+FGMP areas, NGO provided an additional cadre dedicated to home-based GMP delivery. The NGO staff reported regular home visits in HGMP+FGMP areas before the pandemic. During COVID-19, CHCPs in both models were redeployed to vaccination, COVID-19 awareness, and surveillance activities; while NGO staff in HGMP+FGMP areas were reassigned entirely to COVID-19 programmes, resulting in the suspension of home-based GMP:

“*During lockdown, we were busy with vaccination and awareness campaigns. GMP was not happening then*.” (CHCP, FGMP, endline).

### Equipment, supplies, and infrastructure

We observed that government-supplied weighing scales and length boards were available in most CCs at baseline. At endline, many of these equipment’s were reported broken or missing, particularly in FGMP-only areas. But there was no government supplied GMP cards throughout the study period. The NGO supplied GMP cards in HGMP+FGMP areas during baseline only, card availability declined after NGO withdrawal at endline:

“*NGO supplied GMP cards, which helped me to plot child weight and give counselling before COVID. But now I have no cards*.” (CHCP, HGMP+FGMP, endline).

We also observed cracked walls, leaking roofs, and uneven floors in most clinics (21 out of 23 CCs) at baseline, hindered GMP service delivery at CCs:

“*The CC wall is cracked and the roof leaks when it rains; I cannot hang the height sticker. The floor is also broken, and I could not place the weight machine here.*” (CHCP, FGMP, Baseline).

Few CHCPs mentioned measuring weight of sick children at the CC courtyard during COVID-19, to maintain social distancing and curb the infection (Table 3).

### Health information and tracking systems

Throughout the study period, CHCPs continued to maintain paper registers and submit daily online reports. The government’s existing system lacked unique child identifiers at both time points, limiting longitudinal child follow-up. In HGMP+FGMP areas, NGO-supported child tracking systems were used at baseline but ceased during lockdown. At endline, no longitudinal child tracking for GMP was reported in either model (Table 3).

“*Supervisors rarely checked or gave us feedback on GMP data during lockdown*.” (CHCP, FGMP, endline).

### Leadership, governance, financing and coordination

At baseline, national GMP policies existed, but operational guidelines were unavailable. In HGMP+FGMP areas, the NGO developed its own GMP guidelines, training materials, and monitoring tools, trained staff, and distributed educational materials and GMP cards through home visits. One NGO supervisor oversaw 15–20 providers, supporting outreach and community engagement. In FGMP-only areas, GMP was clinic-based with limited promotion. NGO and government services operated largely in parallel in HGMP+FGMP sites, with minimal formal coordination:

“*We were working in the same catchment area, but there was no structured platform to review progress together. Everyone followed their own schedule and planning*.” (NGO Programme Manager, HGMP+FGMP, baseline)

At endline, NGO-supported monitoring mechanisms in HGMP+FGMP areas (spot checks, reporting, feedback loops) declined rapidly after withdrawal. Supervision at CCs was largely administrative with limited technical feedback in both models. CC providers reported limited supervision, coordination, and accountability, contrasting GMP with better-monitored services like immunization and COVID-19 response. Routine supervision was weak, particularly in FGMP-only areas, and data were rarely used for decision-making:

“*We send monthly reports, but we don’t get feedback or discuss the data. It’s more like form filling*.” (CHCP, FGMP-only area, endline).

“*There is lack of monitoring and coordination for GMP and nutrition service at the community clinic; unlike other services (immunization and COVID), which have strict monitoring*.

*Therefore, CC staff does not prioritize GMP activities.”* (National program officer, endline)

During COVID-19, neither government nor NGO had dedicated budgets for GMP programmes. The donor of the NGO withdrew funds from the GMP programme and redirected it to COVID-19 prevention programme. GoB also reallocated budgetary allocations towards COVID-19 prevention programmes (Table 3).

## Discussion

GMP programmes in rural Bangladesh demonstrated limited resilience during the COVID-19 health system shock. Prior to covid-19, HGMP+FGMP areas had initially achieved broader support through routine home visits, EPI integration, GMP card distribution, and caregiver exposure to growth charts, resulting in higher caregiver engagement and service coverage compared with FGMP-only areas. FGMP-only clinics primarily measured child weight, with minimal length/height assessment, no counselling, and no child tracking systems. Across both models, common barriers included workforce shortages, high provider workloads, limited refresher training, weak supervision, equipment and infrastructure gaps, low caregiver awareness, and limited government–NGO coordination^24^. During the pandemic, these challenges intensified, leading to suspension of NGO-led home visits in HGMP+FGMP areas, partial disruption of FGMP clinic services, staff redeployment to COVID-19 duties, supply chain interruptions, and reduced caregiver engagement.

These study findings are consistent with evidence from other LMICs, where child nutrition programmes have been shown to be highly vulnerable to health system shocks^25–27^. Pandemics and crises disrupt routine growth monitoring^26^ ^28–32^, particularly when service delivery relies heavily on NGOs operating within fragile government health systems^24^ ^33^ ^34^. Integrated or home-based models, such as HGMP+FGMP, can improve coverage and caregiver engagement; however, benefits are not sustained without formal institutional support, structured supervision, and contingency mechanisms^1^ ^4^ ^6^ ^31^ ^35–37^. Few LMICs demonstrated resilience in GMP implementation during COVID-19^35^ ^38^. Even where resilience was observed, service quality and coverage were often suboptimal compared with pre-pandemic levels^38^. Experiences from Afghanistan, Democratic Republic of the Congo, Ghana, Sierra Leone, and Ethiopia similarly showed that home-based and integrated nutrition services improved continuity and caregiver engagement, but only where strong coordination, supervisory support, and flexible financing were in place ^39–43^. The reliance on NGO staff and parallel monitoring systems highlights the fragility of government-led GMP programmes, while the absence of shock-resilient policies contributed to service suspension during the pandemic ^33^ ^44^ ^45^. These findings underscore a key lesson for other LMICs: sustainable GMP implementation requires formal integration within government systems rather than dependence on parallel NGO-driven structures.

The limited resilience observed in Bangladeshi GMP programmes reflects systemic weaknesses within the country’s health system across all WHO building blocks rather than isolated implementation gaps. Service delivery was inconsistent and often reduced to weight measurement with minimal counselling, limiting GMP’s preventive and promotive function^46^. Evidence from HGMP+FGMP programmes suggests that structured home visits, community-based child tracking, caregiver engagement with growth charts, and timely referral pathways can enhance programme reach and continuity by shifting GMP from episodic measurement to longitudinal child monitoring^5–7^. Workforce constraints, including high workloads, reliance on NGO capacity, limited refresher training, and rapid redeployment during crises, underscore the absence of adaptive human resource management^47^. Health information systems prioritised routine reporting over actionable use; diminished supervision and feedback during the pandemic further weakened data-driven decision-making^48^. The discontinuation of NGO-supported child tracking removed a critical follow-up mechanism, illustrating the risks of parallel systems not embedded within government structures^49^. Persistent gaps in equipment supply and poor community clinic infrastructure reduced the feasibility of maintaining quality GMP services during crises^7^ ^10^. Weak leadership and governance, marked by limited government–NGO coordination and the absence of shock-resilience guidelines, further constrained adaptive responses and contributed to programme inactivity^49^ ^50^. Inadequate and inflexible financing enabled rapid diversion of resources to emergency response, compromising essential nutrition service delivery during crises^45^. Collectively, these findings indicate that GMP resilience requires institutionalisation within routine government systems, strengthened workforce capacity, integrated data and coordination mechanisms, protected financing, and explicit emergency preparedness during shocks^34^ ^51^ ^52^. Lessons from other LMICs demonstrate that embedding community-based nutrition interventions into routine health systems, combined with clear supervision and rapid resource mobilisation during crises, can sustain service delivery under shocks^53^. These lessons are directly applicable to other LMICs with parallel NGO-government nutrition systems: structured home visits, routine child tracking, caregiver counselling, and integration with broader primary care services can improve resilience during shocks. Establishing formal coordination platforms, emergency SOPs, and dedicated resources can help other countries maintain essential child health services in crises.

Our study had several limitations. The HGMP programme stopped completely and FGMP services were halted during the pandemic, limiting the original plan to assess programme impact on child health and nutritional status. Endline facility observations were not possible due to COVID-19 restrictions, and reliance on self-reported data may introduce recall bias. The inability to observe services during lockdown limits our understanding of real-time adaptation efforts. The sample was relatively small and not nationally representative, limiting generalisability of the study findings. However, COVID-19 offered a unique opportunity to uncover vulnerabilities in both government and NGO systems implementing GMP. The study’s strengths include the inclusion of multiple stakeholders across both programme models, longitudinal assessment from baseline to post-pandemic endline, and triangulation of interviews with facility records and NGO reports, providing a comprehensive understanding of constraints, facilitators, and vulnerabilities using an established framework of health system building blocks.

To strengthen GMP resilience, policies should formalize government–NGO coordination, institutionalize community-based outreach, maintain essential equipment, provide structured supervision and feedback, and allocate dedicated funding and human resources^52^. Countries with similar health system structures can adopt these recommendations, tailoring implementation to local workforce capacity, service delivery contexts, and lessons from other LMICs that successfully maintained child nutrition services during crises. Embedding facilitators observed in HGMP+FGMP programme such as structured home visits, caregiver counselling, and growth chart use into routine government systems could enhance coverage, engagement and continuity during crises^4^. Contingency planning for health system shocks is essential to prevent service disruptions and ensure routine child growth monitoring remains accessible during crises^34^ ^54^.

Strengthening institutional ownership, workforce capacity, coordination, and emergency preparedness is essential to ensure continuity of GMP services during future health system shocks. Other LMICs with similar service delivery structures could apply these lessons when designing or sustaining GMP programmes to improve resilience during health system shocks, highlighting the global relevance of the Bangladeshi experience.

Future research should quantify the impact of service disruptions on child growth outcomes, assess system-level preparedness for emergencies, and evaluate strategies to improve GMP resilience and sustainability. Studies could explore digital monitoring, remote counselling, and hybrid facility–home-based models, examining their ability to maintain service continuity, caregiver engagement, and workforce effectiveness under different types of system shocks.

Comparative studies across countries could further illuminate best practices and contextual adaptations for resilient GMP programmes, providing transferable lessons for LMIC settings.

## Data Availability

No data are available for sharing. Due to its sensitive nature, the interview data remain confidential.

## Acknowledgements

The study co-authors acknowledged the use of ChatGPT 3.5 (free version), developed by OpenAI, to copyedit the English language for grammar. The study team also acknowledges the role of two qualitative data collectors and moderators, Ms. Ishrat Jahan and Dr. Ziaul Islam, for their contribution to qualitative data collection. We also acknowledge all the interview participants for their time and valuable insights. We thank Jocalyn Clark for her comments on an earlier version of this manuscript.

## Authors’ contributions

M.H. designed the study, conducted the data analysis, interpreted the results, and wrote the manuscript. M.H., F.Z., T.H., T.A., U.A., and P.A. contributed to the study design, results interpretation, intellectual content of the manuscript, and edited the manuscript. All authors read and approved the final manuscript.

## Disclaimer

The funders had no role in the study design, data collection, analysis, or interpretation. The research was conducted independently of the funders, and the views expressed in this protocol are those of the authors.

## Patient consent for publication

Not required.

## Patient and public involvement (PPI)

Patients were not involved in the design or conduct of the study. At the initial stages of the study, we established a technical interest group (TIG) comprising members from government, development partners, NGOs, academia and research institutes. This group has informed all stages of the project, including research and intervention design, methodology and the format and manner of communication to participants. We shared study findings with study participants and key stakeholders in a national dissemination.

