## Supplementary material for "Were Bangladesh’s Growth Monitoring and Promotion (GMP) programmes resilient during a health system crisis? A qualitative study"

**Appendix**

**Types and number of interviews conducted**

At **baseline (n = 48)**:

- 4 key informant interviews (KIIs) with government officials (National nutrition program, national community clinic based health care program)
- 8 KIIs with NGO program managers (4) and health workers (4)
- 20 in-depth interviews (IDIs) with caregivers (11 HGMP+FGMP, 9 FGMP)
- 16 IDIs with community clinic (CC) service providers (10 HGMP+FGMP, 6 FGMP)
- Observations at 23 CCs (13 HGMP+FGMP, 10 FGMP) and 5 immunization clinics (HGMP+FGMP area)

At **endline (n = 36)**:

- 4 KIIs with government officials (National nutrition program (2), national community clinic based health care program (2))
- 2 KIIs with NGO program manager (1) and health workers (1)
- 15 IDIs with caregivers (8 HGMP, 7 FGMP)
- 15 IDIs with CC providers (8 HGMP, 7 FGMP)
- No CC observations were conducted at endline

**Consolidated criteria for reporting qualitative studies (COREQ): 32-item checklist**

| **No. Item** | **Guide questions/description** | **Reported on Page #** |
| --- | --- | --- |
| **Domain 1: Research team and reﬂexivity** | | |
| *Personal Characteristics* |  |  |
| 1. Interviewer/facilitator | Which author/s conducted the interview or focus group? | 9 |
| 2. Credentials | What were the researcher’s credentials? E.g. PhD, MD | 1 |
| 3. Occupation | What was their occupation at the time of the study? | 9 |
| 4. Gender | Was the researcher male or female? | 9 |
| 5. Experience and training | What experience or training did the researcher have? | 9 |
| *Relationship with participants* |  |  |
| 6. Relationship established | Was a relationship established prior to study commencement? | 9 |
| 7. Participant knowledge of the interviewer | What did the participants know about the researcher? e.g. personal goals, reasons for doing the research | 12 |
| 8. Interviewer characteristics | What characteristics were reported about the interviewer/facilitator? e.g. Bias, assumptions, reasons and interests in the research topic | 11 |
| **Domain 2: study design** | | |
| *Theoretical framework* |  |  |
| 9. Methodological orientation and Theory | What methodological orientation was stated to underpin the study? e.g. grounded theory, discourse analysis, ethnography, phenomenology, content analysis | 9-11 |
| *Participant selection* |  |  |
| 10. Sampling | How were participants selected? e.g. purposive, convenience, consecutive, snowball | 8 |
| 11. Method of approach | How were participants approached? e.g. face-to-face, telephone, mail, email | 9 |
| 12. Sample size | How many participants were in the study? | 8 |
| 13. Non-participation | How many people refused to participate or dropped out? Reasons? | 9 |
| *Setting* |  |  |
| 14. Setting of data collection | Where was the data collected? e.g. home, clinic, workplace | 9 |
| 15. Presence of non-participants | Was anyone else present besides the participants and researchers? | 9 |
| 16. Description of sample | What are the important characteristics of the sample? e.g. demographic data, date | 12 |
| *Data collection* |  |  |
| 17. Interview guide | Were questions, prompts, guides provided by the authors? Was it pilot tested? | 9 |
| 18. Repeat interviews | Were repeat interviews carried out? If yes, how many? | 9 |
| 19. Audio/visual recording | Did the research use audio or visual recording to collect the data? | 9 |
| 20. Field notes | Were ﬁeld notes made during and/or after the interview or focus group? | 9 |
| 21. Duration | What was the duration of the interviews or focus group? | 9 |
| 22. Data saturation | Was data saturation discussed? | 9 |
| 23. Transcripts returned | Were transcripts returned to participants for comment and/or correction? | 9 |
| **Domain 3: analysis and ﬁndings** | | |
| *Data analysis* |  |  |
| 24. Number of data coders | How many data coders coded the data? | 9 |
| 25. Description of the coding tree | Did authors provide a description of the coding tree? | 9-10 |
| 26. Derivation of themes | Were themes identiﬁed in advance or derived from the data? | 9-11 |
| 27. Software | What software, if applicable, was used to manage the data? | N/A |
| 28. Participant checking | Did participants provide feedback on the ﬁndings? | N/A |
| *Reporting* |  |  |
| 29. Quotations presented | Were participant quotations presented to illustrate the themes/ﬁndings? Was each quotation identiﬁed? e.g. participant number | 14-19 |
| 30. Data and ﬁndings consistent | Was there consistency between the data presented and the ﬁndings? | 14-19 |
| 31. Clarity of major themes | Were major themes clearly presented in the ﬁndings? | 14-19 |
| 32. Clarity of minor themes | Is there a description of diverse cases or discussion of minor themes? | 14-19 |

Developed from: Tong A, Sainsbury P, Craig J. Consolidated criteria for reporting qualitative research (COREQ): a 32-item checklist for interviews and focus groups. *International Journal for Quality in Health Care*. 2007. Volume 19, Number 6: pp. 349 – 357

**Supplementary Table 1. Socio-demographic characteristics of child caregivers**

| **Parameter** | **Baseline** | | **Endline** | |
| --- | --- | --- | --- | --- |
|  | **HGMP+FGMP (n=11)** | **FGMP (n=9)** | **HGMP+FGMP (n=8)** | **FGMP (n=7)** |
| Mother’s age (years), mean (SD) | 25 (6) | 24 (4) | 26 (4) | 25 (4) |
| Mother’s years of education, mean (SD) | 7 (5) | 7 (3) | 9 (5) | 9 (3) |
| Mother’s occupation, housewife, n (%) | 11 (100%) | 9 (100%) | 8 (100%) | 7 (100%) |
| Father’s occupation, n (%) | | | | |
| Service holders | 2 (18%) | 2 (22%) | 4 (50%) | 3 (43%) |
| Day labourer | 5 (45%) | 4 (44%) | 3 (38%) | 2 (29%) |
| Small business | 3 (27%) | 3 (33%) | 0 (0%) | 2 (29%) |
| Unemployed | 1 (9%) | 0 (0%) | 1 (13%) | 0 (0%) |
| Family members per household, mean (SD) | 5 (2) | 5 (1) | 5 (2) | 4 (1) |
| Monthly household income (BDT), mean (SD) | 11,600 (9,192) | 9,944 (6,858) | 13,000 (8,332) | 9,571 (5,533) |
| Monthly household income (USD), mean (SD) | 95 (75) | 81 (56) | 106 (68) | 78 (45) |
| Child age in months, mean (SD) | 11 (3) | 10 (2) | 22 (2) | 22 (4) |

BDT: Bangladeshi Taka; SD: standard deviation; USD: United States Dollar.

**Supplementary Table 2. Characteristics of community clinic healthcare providers**

| **Parameter** | **Baseline** | | **Endline** | |
| --- | --- | --- | --- | --- |
|  | **Baseline HGMP+FGMP (n=10)** | **Baseline FGMP (n=6)** | **Endline HGMP+FGMP (n=8)** | **Endline FGMP (n=7)** |
| Age in years, mean (SD) | 33 (3) | 32 (5) | 34 (3) | 32 (3) |
| Years of education, mean (SD) | 17 (1) | 15 (2) | 15 (2) | 15 (2) |
| Years of experience at CC, mean (SD) | 8 (1) | 8 (0) | 9 (2) | 9 (2) |
| Gender (Female/Male) | 6/4 | 5/1 | 4/4 | 6/1 |

SD: standard deviation.

**Study tools**

**Interview guides aligned to the WHO’s Health System Building Blocks Framework**

This appendix presents the qualitative interview guides used for key informant interviews (KIIs) with government and NGO service providers and in-depth interviews (IDIs) with mothers/caregivers. The guides are explicitly aligned with the World Health Organization (WHO) Health System Building Blocks framework and were designed to capture facilitators, barriers, and adaptations related to growth monitoring and promotion (GMP) service delivery before and during the COVID-19 health system shock.

The six WHO health system building blocks: service delivery; health workforce; health information systems; access to essential medicines, equipment, and infrastructure; leadership and governance; and financing provided an operational and practice-oriented lens to assess health system resilience. Questions were flexibly applied to allow respondents to reflect on routine implementation and changes during the COVID-19 period.

**Key Informant Interview Guide: Government and NGO GMP Service Providers**

**Instruction for the interviewer:** Obtain written informed consent prior to the interview using the approved consent form. Use probes as needed based on the respondent’s role.

*1. Service delivery (WHO building block: Service delivery)*

- Please describe your role and responsibilities related to GMP for children under two years of age.
- Where and how are GMP services usually delivered (community clinic, home visits, EPI clinics)?
- How often are GMP activities conducted, and what services are included (measurement, counseling, referral)?
- How did GMP service delivery change during the COVID-19 period?

*[Purpose:* To assess service availability, continuity, coverage, and adaptations before and during COVID-19.]

*2. Health workforce (WHO building block: Health workforce)*

- Have you received any training related to GMP or child nutrition? When was the most recent training?
- What skills or knowledge did the training focus on (measurement, growth charts, counseling)?
- How were staff roles, workloads, or assignments affected during COVID-19?

*[Purpose:* To explore workforce capacity, training, workload, and redeployment during the health system shock.]

*3. Equipment, supplies, and infrastructure (WHO building block: Access to essential medicines, equipment, and infrastructure)*

- What equipment and supplies are available for GMP (scales, length boards, GMP cards)?
- Are there challenges related to equipment functionality or clinic infrastructure?
- Did availability of equipment or supplies change during COVID-19?

*[Purpose:* To document material and infrastructural facilitators and constraints affecting GMP quality and continuity.]

*4. Health information systems (WHO building block: Health information systems)*

- How do you record and report GMP data?
- Do you use growth charts or registers for tracking children over time?
- How is GMP data supervised or reviewed, and did this change during COVID-19?

*[Purpose:* To assess data collection, use, supervision, and continuity of child tracking systems.]

*5. Leadership and governance (WHO building block: Leadership and governance)*

- Are there any written guidelines or SOPs for GMP or child nutrition services?
- How do government and NGO actors coordinate GMP activities?
- Were there any specific instructions or guidance for GMP service delivery during COVID-19?

*[Purpose:* To examine policy guidance, coordination mechanisms, and crisis preparedness.]

*6. Financing (WHO building block: Financing)*

- Are there dedicated resources or budgets for GMP activities?
- How are GMP activities financed at facility or programme level?
- Did funding priorities change during COVID-19, and how did this affect GMP?

*[Purpose:* To explore financial support, flexibility, and resource diversion during the pandemic.]

*7. Perceived barriers, facilitators, and recommendations (cross-cutting across building blocks)*

- What were the main barriers to GMP service delivery before and during COVID-19?
- What factors helped GMP services function when they did?
- What changes are most urgently needed to strengthen GMP resilience in future crises?

**In-Depth Interview Guide: Mothers/Caregivers of Children Under Two Years**

**Instruction for the interviewer:** Obtain written informed consent prior to the interview using the approved consent form.

1. Service access and utilization (WHO building block: Service delivery)

- Please describe your experience with your child’s weight or length measurement.
- Where did the service take place, and how often was it done?
- Did your access to GMP services change during COVID-19?

*[Purpose:* To capture caregiver-reported access, continuity, and utilization of GMP services.]

2. Counseling and interaction with providers (WHO building block: Health workforce & service delivery)

- Did anyone explain your child’s growth or show you a growth chart?
- Did you receive advice on feeding or caring for your child?
- Were you able to follow the advice given?

*[Purpose:* To understand counseling quality and caregiver engagement.]

3. Understanding and use of growth information (WHO building block: Health information systems)

- Do you understand what your child’s weight or length means for their health?
- What do you think might happen if a child does not grow well?

*[Purpose:* To assess how growth information is communicated and understood at household level.]

4. Barriers to access (WHO building block: Cross-cutting)

- What difficulties do you face in accessing GMP or child nutrition services?
- Were these difficulties different during the COVID-19 period?

*[Purpose:* To identify demand-side barriers interacting with system-level constraints.]

5. Caregiver suggestions (WHO building block: Cross-cutting / governance)

- What would you suggest to improve growth monitoring and child nutrition services for mothers like you?

**Community clinic Observation Checklist**

**Instruction for the interviewer:** Non-participant observation approach will be followed at all study community clinics for capturing quality of GMP care and other nutrition services

Date of observation-----------------------------------

Place of observation--- ------------------------------

Time start------------------ Time end--------------------

Name of observer----------------------------------------

| **Indicators** | **Observation finding and code** | **Observation notes** |
| --- | --- | --- |
| **1.Waiting area** | **Seating arrangement**   - Adequate--------------1 - Not adequate---------2 |  |
|  | **Cleanliness**   - Very good------------1 - Good------------------2 - Not good-------------3 |  |
|  | **Light-air-ventilation**   - Very good------------1 - Good------------------2 - Not good-------------3 |  |
|  | **Toilet**   - Good with water supply------1 - Average condition—2 - Not good--------------3 |  |
| 2.**Waiting time** (approx) in minute per child | - ------------------ (minute) |  |
| 3.Did the provider exchange **welcome greetings** with the mother/caregiver | - Yes---------------------1  - No----------------------2 |  |
| 4**.Supplies**  Length board | Available in good-condition-1  Available but not in good condition------------------------2  Not available-------------------3 |  |
| Weight scale | Available in good-condition-1  Available but not in good condition------------------------2  Not available-------------------3 |  |
| Growth chart/card | Available & enough-----------1  Available but not enough-----2  Not available-------------------3 |  |
| 5. **GMP service provided/received** | | |
| a. Child’s weight measurement | Correctly measured-----------1  Incorrectly measured---------2  Average care-------------------3 |  |
| b. Child’s length measurement | Correctly measured-----------1  Incorrectly measured---------2  Average care-------------------3 |  |
| c. How was GMP data recorded? | Recorded in growth chart correctly-------------------------1  Recorded in growth chart but incorrectly-----------------------2  Recorded in register----------3  Not recorded-------------------4 |  |
| d. Did the mother carry growth card? | Yes---------------------------1  No----------------------------2  Lost---------------------------3  Forgot to carry--------------4 |  |
| e. Did the provider explain GMP finding to mother? | Explained adequately--------1  Explained but not enough---2  Not explained-----------------3 |  |
| f. Did the provider ask mother for any other co-morbidity? | Yes----------------------------1  No---------------------------- 2 |  |
| 6. Did provider examine the baby for co-morbidities? | Yes----------------------------1  No---------------------------- 2 |  |
| 7. Did the provider give counselling based on child growth? | Yes, enough----------------1  Yes, but not enough------- 2  Not done---------------------3 |  |
| 8. Did the provider refer the child at the upazilla health complex (UHC)/ nearest hospital? | Yes----------------------------1  No---------------------------- 2  Not required------------------3 |  |
| 9. Did the provider give date for next follow –up? | Yes----------------------------1  No-----------------------------2 |  |
| 10.Total time spent in GMP per child (Length, weight, counselling, growth chart filling) | -----------------------(minute) |  |
| 11. Who usually provide GMP? | CHCP--------------------------1  HA-----------------------------2  FWA---------------------------3  NGO staff-------------------- 4 |  |
| 12. Who assists in GMP? | HA-----------------------------1  FWA---------------------------2  NGO staff-------------------- 3  None----------------------------4 |  |
